# Clinical efficacy landscaping in genetic obesity: A meta-analysis in Prader Willi syndrome (PWS)

**DOI:** 10.1101/2024.08.02.24311335

**Authors:** Manish Sarkar, Henning von Horsten, Dimitrije Milunov, Nathalie Barreto Lefebvre, Soham Saha

**Author notes:** Corresponding author: *Soham Saha, Medinsights SAS, Paris, France, Email:.

## Abstract

Genetic obesity such as Prader Willi syndrome (PWS) is a multifaceted condition influenced by various elements, including genetic predisposition, environmental factors, and behavioral components. This meta-analysis explored the reported efficacy of therapeutics in clinical trials for PWS by evaluating clinical endpoints reached in the course of the study and the adverse events observed for each. We looked at GLP1 receptor mediated and non-GLP1 receptor mediated therapies and compared their performance in study arms across time and standard of care. In addition, we estimated the present market shares across different obesity and diabetes assets which have been tested against PWS. In conclusion, the study points to two key readouts. First, existing obesity and diabetes assets are not effective for genetic obesity such as PWS. The unmet needs remain high. Second, the markets for obesity and diabetes are in a stage of expansion. A collaborative approach to therapy development for genetic obesity is required to improve the quality of life for affected individuals.

## Introduction

Approximately 7,000 rare diseases affect 4% of the world population of which 72% of the cases are genetic and 70% are pediatric, equivalent to a conservative estimate of 300 million people worldwide (1). Less than 6% of these diseases have an approved therapeutic treatment against them (2), thus underscoring significant gaps in addressing the healthcare needs of those affected. Rare diseases are therapeutic areas with very high unmet needs and repositioning drugs for new indications is a more time-efficient and cost-effective approach compared to developing novel orphan drugs. This method boasts higher success rates, substantially mitigating the risks associated with drug development for rare diseases. The Orphan Drug Designation program takes into account the drugs (small molecules and biologics) which can be applied for treatment with safety and high efficacy against rare diseases which affect less than 200,000 people in the US. This kind of approval follows standard regulatory requirements and pipeline to obtain the status of investigational use. The developers or sponsors of these kinds of drugs must ensure that the drug to be used in the target indication should be safe and efficient backed by well conducted studies.

Genetic obesity poses a considerable challenge in medical research and clinical practice due to its intricate causes and complex traits making it a prime example of multifactorial obesity. This condition is shaped by a mix of genetic tendencies, environmental influences, and behavioral factors, making it a prime example of multifactorial obesity. The importance of clinical effectiveness in genetic obesity is paramount, as it directly affects the development of effective treatments and the overall management of the condition (**Table 1**). The discovery of molecular genetic causes for abnormal weight gain has greatly advanced the understanding and management of obesity, especially in children, leading to the creation of anti-obesity medications and clinical guidelines that address a long-standing gap in care (3).

**Table 1:** Types of genetic obesity indications and genes implicated in them (extracted from human phenotype ontology)

| Disease ID | Disease Name | Gene Symbols |
| --- | --- | --- |
| OMIM:615812 | Abdominal obesity-metabolic syndrome 3 (AOMS3); central obesity; type 2 diabetes; hypertension; early-onset coronary artery disease | DYRK1B |
| OMIM:219080 | ACTH-independent macronodular adrenal hyperplasia | ARMC5, GNAS |
| OMIM:615954 | ACTH-independent macronodular adrenal hyperplasia 2 (AIMAH2) | ARMC5 |
| OMIM:219090 | ACTH-secreting pituitary adenoma; Pituitary Cushing disease | AIP, USP8 |
| OMIM:202155 | Adrenal Hypoplasia, Cytomegalic Type | NR0B1 |
| OMIM:218030 | Apparent Mineralocorticoid Excess 1 | HSD11B2 |
| OMIM:613546 | Aromatase Deficiency | CYP19A1 |
| OMIM:177735 | Autosomal Dominant Pseudohypoaldosteronism Type I | NR3C2 |
| OMIM:604271 | Autosomal idiopathic short stature | GHR |
| OMIM:613192 | Autosomal Recessive Mental Retardation 13 | METTL23, CRADD, NSUN2, FMN2, CLIP1, ZC3H14, PRSS12, LINS1, NDST1, CC2D1A, FBXO31, TRAPPC9, CRBN, GRIK2, SLC6A17, TECR, MAN1B1, TUSC3, ST3GAL3, EDC3, MED13L, KIAA1033, MED23 |
| OMIM:614202 | Autosomal Recessive Mental Retardation 15 | METTL23, CRADD, NSUN2, FMN2, CLIP1, ZC3H14, PRSS12, LINS1, NDST1, CC2D1A, FBXO31, CRBN, TRAPPC9, GRIK2, SLC6A17, TECR, MAN1B1, TUSC3, ST3GAL3, EDC3, MED13L, KIAA1033, MED23 |
| OMIM:264350 | Autosomal Recessive Pseudohypoaldosteronism Type I | SCNN1A, SCNN1B, SCNN1G |
| OMIM:615987 | Bardet-Biedl Syndrome 10 | MKKS, NPHP1, BBIP1, ARL6, BBS1, BBS2, BBS4, BBS9, TTC8, BBS12, TRIM32, BBS7, IFT27, IFT172, WDPCP, SDCCAG8, MKS1, BBS5, CEP290, LZTFL1, BBS10 |
| OMIM:615988 | Bardet-Biedl Syndrome 11 (BBS11) | MKKS, NPHP1, BBIP1, ARL6, BBS1, BBS2, BBS4, BBS9, TTC8, BBS12, TRIM32, BBS7, IFT27, IFT172, WDPCP, SDCCAG8, MKS1, BBS5, CEP290, LZTFL1, BBS10 |
| OMIM:615989 | Bardet-Biedl Syndrome 12 | MKKS, NPHP1, BBIP1, ARL6, BBS1, BBS2, BBS12, BBS4, BBS9, TTC8, TRIM32, BBS7, IFT27, IFT172, WDPCP, SDCCAG8, MKS1, BBS5, CEP290, LZTFL1, BBS10 |
| OMIM:615990 | Bardet-Biedl Syndrome 13 | MKKS, NPHP1, BBIP1, ARL6, BBS1, BBS2, BBS9, BBS4, TTC8, BBS12, TRIM32, BBS7, IFT27, IFT172, WDPCP, SDCCAG8, MKS1, BBS5, CEP290, LZTFL1, BBS10 |
| OMIM:615991 | Bardet-Biedl Syndrome 14 | MKKS, NPHP1, BBIP1, ARL6, BBS1, BBS2, BBS4, BBS9, TTC8, BBS12, TRIM32, BBS7, IFT27, |
|  |  | IFT172, WDPCP, SDCCAG8, MKS1, CEP290, BBS5, LZTFL1, BBS10 |
| OMIM:615993 | Bardet-Biedl Syndrome 16 | MKKS, NPHP1, BBIP1, ARL6, BBS1, BBS2, BBS4, BBS9, TTC8, BBS12, TRIM32, BBS7, IFT27, IFT172, WDPCP, SDCCAG8, MKS1, BBS5, CEP290, LZTFL1, BBS10 |
| OMIM:615995 | Bardet-Biedl Syndrome 18 | MKKS, NPHP1, BBIP1, ARL6, BBS1, BBS2, BBS4, BBS9, TTC8, BBS12, TRIM32, BBS7, IFT27, IFT172, WDPCP, SDCCAG8, MKS1, BBS5, CEP290, LZTFL1, BBS10 |
| OMIM:615996 | Bardet-Biedl Syndrome 19 | MKKS, NPHP1, BBIP1, ARL6, BBS1, BBS2, BBS4, BBS9, TTC8, BBS12, TRIM32, IFT27, BBS7, IFT172, WDPCP, SDCCAG8, MKS1, BBS5, CEP290, LZTFL1, BBS10 |
| OMIM:615981 | Bardet-Biedl Syndrome 2 | MKKS, NPHP1, BBIP1, ARL6, BBS1, BBS2, BBS4, BBS9, TTC8, BBS12, TRIM32, BBS7, IFT27, IFT172, WDPCP, SDCCAG8, MKS1, BBS5, CEP290, LZTFL1, BBS10 |
| OMIM:615982 | Bardet-Biedl Syndrome 4 | MKKS, NPHP1, BBIP1, ARL6, BBS1, BBS2, BBS4, BBS9, TTC8, BBS12, TRIM32, BBS7, IFT27, IFT172, WDPCP, SDCCAG8, MKS1, BBS5, CEP290, LZTFL1, BBS10 |
| OMIM:615983 | Bardet-Biedl Syndrome 5 | MKKS, NPHP1, BBIP1, ARL6, BBS1, BBS2, BBS4, BBS9, TTC8, BBS12, TRIM32, BBS7, IFT27, IFT172, WDPCP, SDCCAG8, MKS1, BBS5, CEP290, LZTFL1, BBS10 |
| OMIM:605231 | Bardet-Biedl Syndrome 6 | MKKS, NPHP1, BBIP1, |
|  |  | ARL6, BBS1, BBS2, BBS4, BBS9, TTC8, BBS12, TRIM32, BBS7, IFT27, IFT172, WDPCP, SDCCAG8, MKS1, BBS5, CEP290, LZTFL1, BBS10 |
| OMIM:615984 | Bardet-Biedl Syndrome 7 | MKKS, NPHP1, BBIP1, ARL6, BBS1, BBS2, BBS4, BBS9, TTC8, BBS12, TRIM32, BBS7, IFT27, IFT172, WDPCP, SDCCAG8, MKS1, BBS5, CEP290, LZTFL1, BBS10 |
| OMIM:615985 | Bardet-Biedl Syndrome 8 | MKKS, NPHP1, BBIP1, ARL6, BBS1, BBS2, TTC8, BBS4, BBS9, BBS12, TRIM32, BBS7, IFT27, IFT172, WDPCP, SDCCAG8, MKS1, BBS5, CEP290, LZTFL1, BBS10 |
| OMIM:615986 | Bardet-Biedl Syndrome 9 | MKKS, NPHP1, BBIP1, ARL6, BBS1, BBS2, BBS9, BBS4, TTC8, BBS12, TRIM32, BBS7, IFT27, IFT172, WDPCP, SDCCAG8, MKS1, BBS5, CEP290, LZTFL1, BBS10 |
| OMIM:602522 | Bartter Syndrome Type 4A | BSND, CLCNKA, CLCNKB |
| OMIM:602025 | Body Mass Index Quantitative Trait Locus 9 (BMIQ9) | MC3R |
| OMIM:300869 | Chromosome Xq27.3-Q28 Duplication Syndrome | FMR1 |
| OMIM:216550 | Cohen Syndrome | VPS13B |
| OMIM:300200 | Congenital Adrenal Hypoplasia | NR0B1 |
| OMIM:214700 | Congenital Secretory Chloride Diarrhea | SLC26A3 |
| OMIM:203400 | Corticosterone Methyloxidase Type I Deficiency | CYP11B2 |
| OMIM:610600 | Corticosterone Methyloxidase Type II Deficiency | CYP11B2 |
| OMIM:604931 | Cortisone Reductase Deficiency 1 | HSD11B1, H6PD |
| OMIM:614662 | Cortisone Reductase Deficiency 2 | HSD11B1, H6PD |
| ORPHANET:189427 | Cushing syndrome due to macronodular adrenal hyperplasia | ARMC5 |
| OMIM:256450 | Familial Hyperinsulinemic Hypoglycemia | ABCC8 |
| OMIM:601820 | Familial Hyperinsulinemic Hypoglycemia 2 | KCNJ11 |
| OMIM:616026 | Fanconi Renotubular Syndrome 4 With Maturity-Onset Diabetes Of The young | HNF1B, HNF4A |
| OMIM:614736 | Glucocorticoid Deficiency 4 | STAR, MRAP, NNT, TXNRD2, MC2R |
| OMIM:262500 | Growth Hormone Insensitivity Syndrome | GHR |
| ORPHANET:30925 | Hereditary Central Diabetes Insipidus | AVP |
| OMIM:610628 | Hypogonadotropic Hypogonadism 4 With Or Without Anosmia | PROK2, PROKR2, SEMA3A, IL17RD, WDR11, KISS1, SOX10, FEZF1, GNRH1, FGF8, GNRHR, ANOS1, TAC3, HS6ST1, CHD7, FGFR1, HESX1, TACR3, FGF17, FLRT3, DUSP6, SPRY4, KISS1R, NSMF |
| OMIM:240900 | Hypoinsulinemic Hypoglycemia With Hemihypertrophy | AKT2 |
| OMIM:613090 | Infantile Bartter Syndrome Type 4B | BSND, CLCNKA, CLCNKB |
| OMIM:245800 | Laurence-Moon Syndrome | PNPLA6 |
| OMIM:614037 | Leukotriene C4 Synthase Deficiency | LTC4S |
| OMIM:615980 | Lipodystrophy, Familial Partial, Type 6 | LIPE |
| OMIM:605309 | Macrocephaly/Autism Syndrome | PTEN |
| OMIM:613375 | Maturity-Onset Diabetes Of The Young Type 11 (Mody11) | BLK, CEL, PDX1, HNF4A, APPL1, KLF11, INS, HNF1A, ABCC8, GCK, PAX4, KCNJ11, NEUROD1 |
| OMIM:613670 | Mental Retardation With Language Impairment And With Or Without Autistic features | FOXP1 |
| OMIM:610156 | Mental Retardation; Truncal Obesity; Retinal Dystrophy; Micropenis | INPP5E |
| OMIM:615418 | Mitochondrial DNA Depletion Syndrome 12 (Cardiomyopathic Type) | SLC25A4, AGK |
| OMIM:614894 | Monocyte And Dendritic Cell Deficiency, Autosomal Recessive;;Irf8 Deficiency, Autosomal Recessive | IRF8 |
| OMIM:615703 | Morbid Obesity And Spermatogenic Failure | CEP19 |
| OMIM:614250 | Narcolepsy 7 | HLA-DRB1, MOG, HCRT, CTSH, P2RY11, ZNF365, TNFSF4, HLA-DQB1 |
| ORPHANET:2073 | Narcolepsy-Cataplexy | HLA-DRB1, HCRT, MOG, CTSH, P2RY11, ZNF365, TNFSF4, HLA-DQB1 |
| OMIM:614962 | Nonsyndromic morbid Obesity 1 | LEP |
| OMIM:614963 | Nonsyndromic morbid Obesity 2 | LEPR |
| OMIM:601665 | Obesity | MC4R, PPARG, SIM1, NR0B2 |
| OMIM:613886 | Obesity; Hyperphagia; Developmental Delay | NTRK2 |
| OMIM:616185 | Ovarian Dysgenesis 4 | MCM9 |
| OMIM:610489 | Pigmented Nodular Adrenocortical Disease, Primary, 1 | PRKAR1A, PDE11A, PDE8B, PRKACA |
| OMIM:610475 | Pigmented Nodular Adrenocortical Disease, Primary, 2 | PRKAR1A, PDE11A, PDE8B, PRKACA |
| OMIM:615830 | Pigmented Nodular Adrenocortical Disease, Primary, 4 | PRKAR1A, PDE11A, PRKACA, PDE8B |
| OMIM:176270 | Prader-Willi Syndrome | NDN, SNRPN |
| OMIM:615547 | Prader-Willi-Like Syndrome | MAGEL2 |
| ORPHANET:189439 | Primary Pigmented Nodular Adrenocortical Disease | PRKAR1A, PDE11A, PDE8B, PRKACA |
| OMIM:614190 | Primary Pigmented Nodular Adrenocortical Disease 3 | PRKAR1A, PDE11A, PDE8B, PRKACA |
| OMIM:609734 | Proopiomelanocortin Deficiency | POMC |
| OMIM:600955 | Proprotein Convertase 1/3 Deficiency | PCSK1 |
| OMIM:603233 | Pseudohypoparathyroidism Type Ib | STX16, GNAS, GNAS-AS1 |
| OMIM:616188 | Retinal Dystrophy And Obesity | RLBP1, SPATA7, ARL6, CERKL, PRPF31, TTC8, PROM1, LRAT, IFT140, BEST1, NEK2, ARL2BP, NR2E3, EYS, MAK, SAG, ABCA4, PDE6A, PDE6G, IMPDH1, OFD1, GUCA1B, NRL, PDE6B, KLHL7, PRPF4, PRPF3, FAM161A, IFT172, KIZ, CDHR1, ZNF408, RBP3, HGSNAT, FSCN2, PRCD, BBS2, RDH12, PRPH2, DHDDS, |
|  |  | ROM1, C2ORF71, RP9, PRPF6, RP1, RP2, RPGR, SEMA4A, C8ORF37, IDH3B, MERTK, PRPF8, TOPORS, USH2A, RPE65, CLRN1, CNGB1, TUB, RGR, CNGA1, SNRNP200, SLC7A14, TULP1, RHO, CA4, CRB1, IMPG2, ZNF513, CRX |
| ORPHANET:791 | Retinitis Pigmentosa | RLBP1, SPATA7, ARL6, CERKL, PRPF31, TTC8, PROM1, LRAT, IFT140, BEST1, NEK2, ARL2BP, NR2E3, EYS, MAK, SAG, ABCA4, PDE6A, PDE6G, IMPDH1, OFD1, GUCA1B, NRL, PDE6B, KLHL7, PRPF4, PRPF3, FAM161A, IFT172, KIZ, CDHR1, ZNF408, RBP3, HGSNAT, FSCN2, PRCD, BBS2, RDH12, PRPH2, DHDDS, ROM1, C2ORF71, RP9, PRPF6, RP1, RP2, RPGR, SEMA4A, C8ORF37, IDH3B, MERTK, PRPF8, TOPORS, USH2A, RPE65, CLRN1, CNGB1, RGR, CNGA1, TUB, SNRNP200, SLC7A14, TULP1, RHO, CA4, CRB1, IMPG2, ZNF513, CRX |
| OMIM:616629 | Senior-Loken Syndrome 9 | WDR19, CEP164, TRAF3IP1, NPHP1, NPHP4, SDCCAG8, NPHP3, CEP290, IQCB1, INVS |
| OMIM:601410 | Transient Neonatal Diabetes Mellitus 1 | ABCC8, HYMAI, KCNJ11, PLAGL1, ZFP57 |
| OMIM:275400 | Trichomegaly With Mental Retardation, Dwarfism, And Pigmentary Degeneration | PNPLA6 |
| OMIM:309585 | Wilson-Turner X-Linked Mental Retardation Syndrome | HDAC8 |
| OMIM:300957 | X linked Mental Retardation | THOC2 |
| OMIM:300582 | X-Linked Idiopathic Short Stature | SHOX |
| OMIM:300354 | X-Linked, Syndromic, Mental Retardation, Cabezas Type (MRXSC) | CUL4B |

Genetic obesity is a polygenic disorder characterized by excessive weight gain due to inherited genetic factors, environmental influences like diet, physical activity and habits. It is often categorized within the broader scope of multifactorial obesity, which encompasses both genetic and non-genetic influences (4). Genetic obesity specifically refers to obesity significantly influenced by genetic factors, although it often interacts with environmental and lifestyle elements. Genetic obesity can be classified based on the genetic mechanisms involved and the presence of other contributing factors:

● Monogenic Obesity: This form is caused by mutations in a single gene. Examples include mutations in the leptin gene, leptin receptor gene, and melanocortin 4 receptor gene (5–8).
● Polygenic Obesity: This is the most common form of genetic obesity, resulting from the combined effect of multiple genes, each contributing a small effect. The exact genetic disorders of its appearance have not been identified yet, demonstrating its polygenic nature (9).
● Syndromic Obesity: This type is associated with other clinical features and is part of a syndrome, such as Prader-Willi syndrome or Bardet-Biedl syndrome (10–12).

In this study, we performed a detailed use case against one of the rare genetic disorders, Prader Willi Syndrome (PWS), with obesity and diabetes as primary clinical features. Obesity and Diabetes are two of the most frequent public health problems that have emerged worldwide from and after 2010 (13). Global obesity is a bigger health crisis than hunger and is a leading reason for fatality and disabilities within the disease space. Historically these were considered as adult diseases, however an increase in weight leading to type 2 diabetes (T2D) has been recorded in children (Age<10) and adolescents (Age: 10-19) (14). Obesity in the United States affects 4 out of 10 people as of present (15). In 2021, close to 40 million people in the United States, which is around 12% of their total population, had diabetes (16). The majority of the diabetes (90-95%) cases are T2D (17). There are two kinds of obesity: (i) Lifestyle obesity (18) and (ii) Genetic obesity (17) comprising of Prader Willi Syndrome, Bardet-Biedl syndrome (BBS), Kleefstra syndrome- EHMT1 mutation, Cohen syndrome-VPS13B/COH1 mutation, 5p13 microdeletion syndrome, CHOPS syndrome-AFF4 mutation, Carpenter syndrome-RAB23 mutation, Alstrom syndrome-ALMS1 mutation, OBHD syndrome-NTRK2 mutation, 16p11.2 deletion, Rubinstein Taybi syndrome-CREBBP mutation, Albright hereditary osteodystrophy associated with GNAS mutation and others (**Table 1**) (19). PWS is a genetic disorder which affects multiple organs and is caused by deletion driven genetic malfunction. This disease is characterized by mental, physical and behavioral problems along with development of hyperphagia i.e., a constant sense of insatiable hunger from an age of around 2 years. Patients with PWS thus have weight gain problems and a wide spectrum of complications of PWS are caused due to obesity (20). The symptoms of PWS vary from newborn to early childhood and adulthood (21). The symptoms present in the infant born with PWS are:

● leading to increased frequency of T2D
● Underdeveloped sex organs
● Growth hormone deficiency
● Cognitive impairment
● Underdeveloped motor responses
● Speech and behavioral problems
● Sleep disorder

PWS arises primarily from the deletion of the paternal 15q.11-13 region in the majority (50-70%) of the cases (PW-del) (**Figure 1a**). A lesser percentage (20-30%) of cases result from maternal uniparental disomy (PW-UPD) due to maternal inheritance of chromosome 15 (**Figure 1b**) and 3% of the patients have paternally contributed genetic imprinting (microdeletions) (**Figure 1c**) (22).

**Figure 1:**
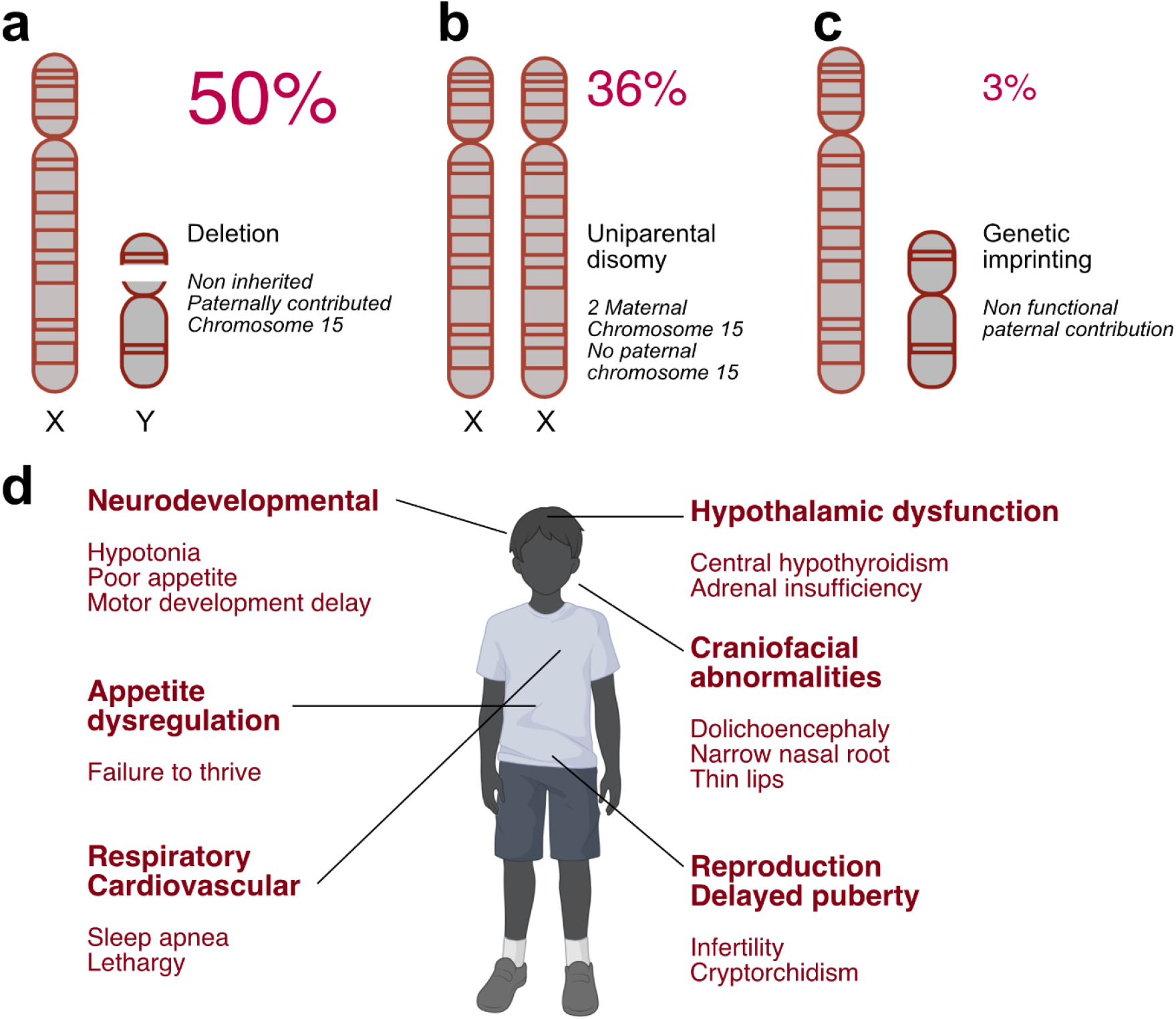
Genetic anomalies in Prader Willi syndrome and key systems impacted. (a) Approximately 50% of PWS cases are due to chromosomal deletion of paternal 15q.11-13 region. (b) Around 36% of all PWS cases are due to uniparental disomy due to maternal inheritance of chromosome 15. (c) A minor proportion of PWS patients are due to genetic imprinting. (d) A infographic showing key systems impacted in PWS and their associated physical features.

The 15q.11-13 chromosomal locus is divided into two regions: (i) critical region (15q11.2-q13.1) and (ii) non critical region. The key genes (23) in the critical region include

● MAGEL2, MKRN3, PWRN1, NDN, SNURF-SNRPN and NPAP1.
● a family of small nucleolar RNA (snoRNA) genes: SNORD116, SNORD107.
● IPW, a long noncoding RNA.

This chromosomal region either directly or indirectly impacts numerous tissues and functions by regulation of genes involved in neuron specific silencing (22), mRNA expression downregulation of proprotein convertase-1 (PCSK1) in the hypothalamus, leading to obesity (19). As a consequence, numerous genes in the critical and non-critical regions have been observed to be differentially expressed in both PW-del (del = paternal deletion) and PW-UPD cases (UPD = maternal uniparental disomy) (24). This study additionally talks about expression changes of specific genes beyond the critical (15q11-q13.1) region which are core to PWS disease progression and pathophysiology. Genes related to neurodevelopmental functions such as PEX10, CTDP1, DHRS1, SNTB2, AKT1S1, etc. show significant upregulation (24). Key cellular functions regulated by the PWS expressed genes are adipogenesis, BDNF regulation of transmission and chronic inflammation mediated by numerous factors (25–27). PWS caused byPW-UPD results in a less severe phenotype compared to PWS caused by PW-del. However, cases of UPD have a higher risk for autism spectrum disorder (ASD) compared to typically developing individuals. Maternal PWS has mitochondrial defects that lead to differential expression of several mitochondria related genes which were identified using DAVID18 analysis, such as NDUFA1, GSTP1, MRPL4, MFN2, NT5M, MINOS1, MTIF2, TIMM17A, TMTC1, SLC25A36, etc. (24).

In summary, PWS leads to a spectrum of clinical manifestations, with appetite dysregulation leading to hyperphagia and obesity being the most prominent and challenging clinical features (**Figure 1d**). Recent developments in the field of obesity drug development have targeted Glucagon-like peptide 1 (GLP1) receptors and have demonstrated promising clinical results (28). However GLP1 receptor agonists (GLP1 RAs) have been found to be ineffective against genetic obesity like PWS (29,30). There remains a high clinical unmet need as far as therapeutics against PWS is concerned. In this study, we have explored key aspects of understanding PWS, in terms of market analysis, competitive intelligence and drug repositioning. Our present study provides a detailed overview covering the following aspects of rare disease evaluation criteria:

1. Competitive intelligence involving clinical trials and patents along with potential competitors in therapeutic development against PWS.
2. Clinical and commercial unmet needs identification and existing drug efficacy landscaping against PWS.
3. Market research and commercial landscaping of the drugs targeting the present standard of care (GLP1 RA) provides us with critical entry points to drug repositioning.

## Methodology

### Data sources

Data was extracted from publicly available information from ClinicalTrials.gov (31), publications discussing the pathophysiology, results of different clinical trials, case reports and annual financial reports of companies of Novo Nordisk, Eli Lilly, Astrazeneca and Sanofi from 2018 to 2023 for GLP1 RAs and Milendo therapeutics, Soleno Therapeutics, Rhythm Pharmaceuticals, Marimar therapeutics and Saniona therapeutics for non GLP 1 modalities in 2023.

### Clinical efficacy landscaping

A meta-analysis study was performed to elucidate different aspects of the clinical trials landscape launched against PWS. Some of the key deliverables evaluated in this study include drug name, mechanism of action, therapy area and clinical phases followed by the sponsors of these clinical trials. The second set of outputs analyzed the primary clinical endpoints of these interventions for PWS trials taking into account GLP1 and non GLP1 mediated benchmarks. For GLP1 targeted therapies, weight loss percentage, BMI moss percentage, glycated hemoglobin concentration decrease and post treatment glycated hemoglobin concentration in the blood, were analyzed as the co primary clinical endpoints. These study took into account outcomes from multiple arms of the clinical trials to analyze the consistency of the intervention efficacy. For non GLP1 mediated interventions, hyperphagic behaviors, weight loss, behavioral and motor responses were evaluated as the primary endpoints. Adverse events were mapped from CT.gov: results section, case reports and clinical trial reports for all the clinical trials analyzed.

### Market analysis

Market analysis of the interventions explored against PWS driven hyperphagia and obesity was performed. The global obesity market size information were extracted from Morgan Stanley Research Estimates(32) between 2022 to 2028 and plotted against the timeline to understand the total market growth of obesity. The global diabetes market size information was extracted from Statista (33) between 2022 to 2028 and plotted against the timeline to understand the total market growth of diabetes. The individual revenues of the interventions were extracted from annual reports of the companies holding market authorization. The proportion of the revenues were calculated in correlation to the total market size of obesity or diabetes medications, to calculate the market share of each drug. The individual EBITDA of each of the companies were extracted from their annual reports. The product specific revenues were calculated as a percentage of the total revenue and used for the determination of product specific EBITDA from companywide EBITDA.

### Clinical Complexities and challenges of PWS

PWS is a complex disorder affecting multiple bodily systems. Individuals with PWS experience a range of symptoms including appetite imbalance, low muscle tone, growth abnormalities, delayed puberty, sleep disturbances, reduced pain sensitivity, neurodevelopmental disorders, musculoskeletal abnormalities and digestive issues (**Figure 1d**). A hallmark characteristic of PWS, distinct from other neurodevelopmental conditions, is a dramatic shift in eating behavior. While infants often struggle with feeding and may fail to thrive, children develop an insatiable appetite. This uncontrolled desire to eat, combined with a slower metabolism, inevitably leads to severe obesity unless strictly managed. The underlying cause of this excessive hunger remains unknown, and there are currently no approved treatments specifically for hyperphagia or other core symptoms of PWS (34).

PWS is characterized by mental, physical and behavioral problems along with development of hyperphagia i.e., a constant sense of insatiable hunger from an age of around 2 years (**Figure 1d**). Patients with PWS thus have weight gain problems and a wide spectrum of complications of PWS are caused due to obesity (20). The symptoms of PWS vary from newborn to early childhood and adulthood (21). The symptoms present in the infant born with PWS are:

● Poor muscle tone - hypotonia
● Reduced sucking reflex
● Abnormal facial features
● Poorly developed genital organs - cryptorchidism in males and underdeveloped clitoris and labia in females
● Late responsiveness and unusual tiredness

The symptoms present from early childhood to adulthood are:

● Hyperphagia and Obesity leading to increased frequency of T2D
● Underdeveloped sex organs
● Growth hormone deficiency
● Cognitive impairment
● Underdeveloped motor responses
● Speech and behavioral problems
● Sleep disorder

### Hyperphagia in PWS

Hyperphagia in PWS is characterized by an overwhelming and persistent craving for food. Individuals with this condition experience intense hunger, obsessive thoughts about food, and difficulty feeling full. They often engage in compulsive food-seeking behaviors and struggle to control their eating. Hyperphagia represents the most severe form of overeating (35).

Hyperphagia in PWS develops in stages. Initially, infants with PWS exhibit low muscle tone, sleepiness, and poor feeding. This phase is often followed by a period of seemingly normal growth (36). However, children with PWS subsequently gain weight rapidly and develop an intense preoccupation with food. Most individuals progress to hyperphagia, characterized by insatiable hunger and an overwhelming urge to eat. Onset typically occurs around age eight, but can vary (36). This excessive eating behavior is likely due to persistent hunger, with temporary satiety only achieved after consuming excessive calories. Individuals with PWS can consume enormous quantities of food at once, often hoarding or eating discarded items. Extreme overeating has led to severe complications, highlighting the lack of normal satiety. While hyperphagia is typically lifelong, its severity can fluctuate (37). Factors such as environmental changes, rather than gender or PWS genetic subtype (38), seem to influence eating behaviors. Importantly, the degree of obesity in PWS is primarily determined by food availability, not the severity of hyperphagia. PWS-associated hyperphagia shares similarities with conditions like binge eating disorder, addiction, and obsessive-compulsive behaviors, particularly the preoccupation with food (39,40). However, it is distinct in its early onset, typically appearing in early childhood, and underlying physiological mechanisms (41). Research suggests a fundamental impairment in satiety regulation in individuals with PWS (**Figure 2**), evidenced by abnormal brain responses to food cues (42). Studies have shown that unlike individuals with binge eating disorder who experience temporary relief from negative emotions after overeating, those with PWS lack this satisfaction and continue to crave food even after excessive consumption (**Figure 2**). Additionally, unlike addictive behaviors which may be driven by a desire for pleasure or reward, hyperphagia in PWS seems to be driven by a more primal need to fulfill a physiological deficit (**Figure 2**).

**Figure 2:**
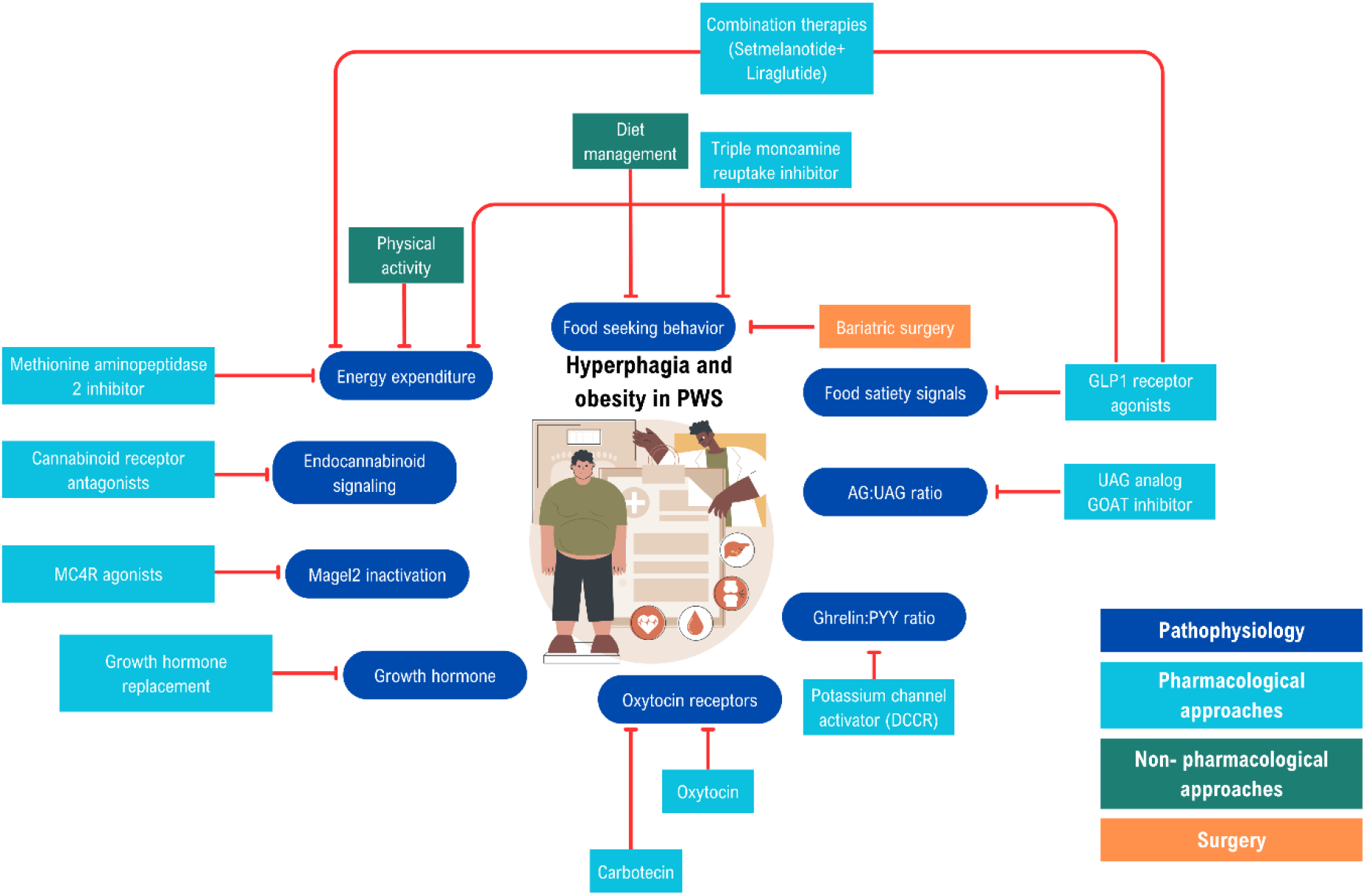
Therapeutic complexity and causative factors of hyperphagia and obesity in PWS. A map showing the different factors influencing hyperphagia and obesity in PWS. The dark blue boxes indicate pathophysiological features, the light blue boxes indicate pharmacological approaches to PWS therapy, the green boxes indicate non-pharmacological approaches and the orange box indicates surgical interventions. Adapted from Tan et al., 2019 (44,45).

For caregivers, managing PWS-related hyperphagia is a relentless and often overwhelming challenge (**Figure 2**). This involves constant surveillance, with caregivers needing to be vigilant about the individual with PWS’s whereabouts and potential access to food. Disruptions to routines, even minor changes to meal schedules or unexpected access to food, can lead to significant behavioral disturbances in individuals with PWS. These disturbances can range from anxiety and frustration to tantrums and even self-injurious behaviors.

Hyperphagia is described as a life-threatening condition that severely impacts quality of life for individuals with PWS and their families. The constant drive to eat can lead to obesity, which in turn increases the risk of developing serious health complications such as heart disease, diabetes, and sleep apnea. The restricted lifestyle imposed by the need for food security can also lead to social isolation and hinder participation in daily activities (35,43).

### Behavioral aggression in PWS

Behavioral aggression in PWS is characterized by intense emotional outbursts that appear disproportionate to the triggering event and are seemingly uncontrollable (46). Aggressive behaviors could be episodic or persistent, and they are a pervasive and debilitating challenge for individuals with PWS. These episodes significantly impact quality of life, rivaling or even surpassing the challenges posed by hyperphagia. In developing children, PWS outbursts have a later onset, greater intensity, and a lifelong course (46).

Temper outbursts in individuals with PWS often involve a predictable pattern of escalating behaviors. These can include repetitive questioning, angry facial expressions, excessive salivation, loud speech, crying, yelling, stomping, slamming, kicking, hitting, lying on the floor, and property damage. The outburst typically begins with emotional displays like crying and vocalizations, rapidly escalating to anger and physical manifestations such as aggression. While early intervention may prevent a full-blown outburst, there’s often a point of no return. These episodes can last from minutes to hours, followed by a period of withdrawal or sleep. The individual may subsequently express remorse or distress (47).

These outbursts are influenced by a complex interplay of factors, including impaired emotional regulation, limited executive function, and potential dysregulation of the autonomic nervous system (47–50). Triggers commonly involve thwarted desires, social violations, perceived unfairness, or changes in routine. Notably, PWS outbursts often precede the onset of hyperphagia, suggesting independent developmental trajectories. One possible mechanism driving the temper outbursts triggered by changes in routine may be a deficit in task-switching ability, a cognitive-related executive function that is an area of particular difficulty in individuals with PWS (51,52). Dysregulation of the autonomic nervous system may be another key mechanism underlying temper outbursts in PWS. The far-reaching consequences of these behaviors extend beyond the individual, impacting families, social interactions, and employment opportunities (46). Understanding the underlying mechanisms of temper outbursts is crucial for developing effective interventions and improving the lives of individuals with PWS.

### Anxiety in PWS

Anxiety is a prevalent issue in PWS. Individuals with PWS often experience excessive worry and tension, frequently centered around routines, meal planning, food security, or specific people or objects. The prospect of change in these areas can significantly trigger anxiety. While there are overlaps between anxiety in PWS and generalized anxiety disorder (GAD), the condition also presents unique characteristics. For example, the intense focus on food-related concerns and specific individuals or items often exceeds the scope of traditional GAD diagnostic criteria (53).

While often associated with food-related concerns and routines, anxiety in PWS extends to various situations, including transitions and changes (52). While its outward manifestations can vary in severity, individuals with PWS frequently exhibit a set of characteristic behaviors indicative of anxiety. These include repetitive questioning, particularly about routines, food, or people; pacing; rapid or loud speech; excessive physical movement; trembling; and compulsive checking behaviors. Those with stronger verbal abilities often express intense worry and feelings of overwhelm. It’s important to note that anxiety and compulsive behaviors often intersect in individuals with PWS, making accurate diagnosis and intervention complex.

Despite the widespread recognition of anxiety by parents and clinicians, research on the prevalence and nature of anxiety in PWS has been inconsistent (54). While some studies have identified moderate to high rates of anxiety, others have underestimated its impact, potentially due to challenges in measurement and definition (55,56). Einfeld et al. found that over 40% of individuals with PWS exhibited elevated anxiety levels, as measured by the Developmental Behavior Checklist (56). Moreover, Feighan et al. reported anxiety as the most common psychiatric diagnosis among adolescents and adults with PWS, affecting nearly 40% of participants (57). These findings underscore the critical importance of addressing anxiety in PWS care. Notably, caregivers consistently rank anxiety as a top concern, emphasizing its detrimental effects on individuals with PWS and their families. According to a 2018 analysis of the PWS Global Patient Registry, nearly half of participants aged 10 and older were reported by caregivers to have significant anxiety symptoms, highlighting the substantial impact of this challenge on the PWS community (58).

Findings suggest that anxiety symptoms are more pronounced in individuals with the UPD genetic subtype compared to those with the deletion subtype. While the exact age of onset remains unclear, anxiety often emerges during preschool or early school years, intensifying during adolescence and early adulthood (59,60). These studies underscore the urgent need for comprehensive research to better understand the complexities of anxiety in PWS, inform effective interventions, and improve the quality of life for individuals affected by this condition.

### Obsessive compulsive behaviors in PWS

Individuals with PWS often exhibit obsessive-compulsive behaviors (OCBs). These patterns typically manifest as rigid adherence to routines, compulsive collecting or hoarding, and an intense need for sameness. Additionally, a persistent drive to acquire and share specific information can be observed, often accompanied by significant distress if prevented. These OCBs can significantly impact daily life and require tailored interventions (61–64).

While previous research indicated high rates of OC-like symptoms in PWS, subsequent studies have shown that these behaviors often differ significantly from classic OCB (63). In a subsequent longitudinal follow-up of over 250 people with PWS, it was found that only 8% of pooled participants met full DSM-5 criteria for diagnosis of obsessive compulsive disorder (OCD). Individuals with PWS frequently exhibit repetitive behaviors, routines, and a strong need for consistency, which can be characterized as OCBs (65). However, unlike OCD, these behaviors often bring pleasure or relief rather than distress. Additionally, the content of obsessions in PWS typically diverges from the typical OCD themes of contamination, harm, or religious doubt. Repetitive questioning is a particularly common OCB in PWS, often causing significant challenges for caregivers.

The onset of OCBs in PWS often appears early in childhood, with some studies indicating higher rates in young children with PWS compared to typically developing peers or children with Down syndrome (66). While research on the relationship between PWS genetic subtypes and OCBs is limited, some studies suggest potential differences (67). It’s crucial to recognize that the diagnostic criteria for OCD may not fully capture the unique characteristics of OCBs in PWS. Further research is needed to better understand the underlying mechanisms and develop targeted interventions for individuals with PWS. OCBs in PWS present with varying degrees of severity and complexity. Nevertheless, a common pattern emerges, characterized by rigid adherence to routines, an obsessive need for information, compulsive collecting, and repetitive actions. These behaviors often manifest as insistence on sameness, excessive questioning or sharing of information, hoarding of objects, and repetitive tasks such as rewriting or rearranging.

### Sleep disorders in PWS

Sleep disturbances are prevalent in individuals with PWS and may contribute to other core symptoms of the disorder. While sleep apnea is often the primary focus of sleep evaluations, a growing body of evidence suggests a broader spectrum of sleep disorders in this population. Excessive daytime sleepiness, in particular, is highly prevalent and persists despite treatment for sleep apnea. While obstructive sleep apnea (OSA) has traditionally been the primary focus, emerging research highlights a more complex sleep phenotype. Genetic, neurological, and hormonal factors converge to create a multifaceted sleep disorder profile in individuals with PWS. Studies at the molecular level have implicated the SNORD116 gene, a key genetic factor in PWS, in regulating sleep-wake cycles, suggesting a direct genetic link to sleep dysfunction (68–70).

Beyond the laboratory, real-world experiences underscore the breadth of sleep issues in PWS (71). Caregivers consistently report a range of sleep problems, including excessive daytime sleepiness (EDS), insomnia, and narcolepsy, in addition to OSA. These challenges extend beyond the physical realm, significantly impacting quality of life, behavior, and cognitive function (72). The intricate relationship between sleep and other PWS symptoms is becoming increasingly evident. Sleep disturbances may contribute to the irritability, behavioral challenges, and cognitive difficulties often observed in individuals with PWS. To fully comprehend and address the complexities of PWS, a comprehensive understanding of sleep disorders is essential. Despite growing recognition, significant gaps persist in our knowledge of the underlying mechanisms and optimal treatment strategies. Further research is imperative to unravel the intricacies of sleep in PWS, ultimately leading to improved patient outcomes.

### Competitive landscaping of therapies against PWS in clinical trials

Prader Willi Syndrome has been targeted by a wide range of therapeutics encompassing a broad spectrum of modalities. Oxytocin, Diazoxide choline, cannabidiol, somatotropin and Carbetocin are the top interventions with the most number of clinical trials (**Figure 3a**). These drugs function by mimicking oxytocin, stimulating growth hormone, blocking NPY-AgRP, and activating cannabinoid receptors (**Figure 3b**). Looking into the entire landscape from a bird view, targeted therapies comprises the highest proportion of the clinical trials followed by hormonal therapy, enzyme replacement therapy and health supplemention (**Figure 3c**). There are 97 interventional clinical trials registered against PWS (31) but there is a lack of GLP1 agonist based monotherapy/combinatorial therapy options. Liraglutide (Saxenda) of Novo Nordisk did not reach the significant efficacy benchmark in obesity related primary endpoints against PWS driven obesity (73). The PWS clinical trials are mostly in Phase 2, followed by Phase 3, Phase 1/2, Phase 2/3, Phase 4 and Phase 1 (**Figure 3d**). This means most of the assets are in late development which has met with developmental blockades due to inefficacy or adverse events. This opens up a huge potential of developing new drugs be it new medicinal product or repositioned drugs against PWS and potentially expand it to the genetic obesity space using basket trial approach. Some of the key players involved in carrying out these clinical trials are Radius Pharmaceuticals, Soleno therapeutics, Essentialis, Novo Nordisk, Saniona and Zafgen (**Figure 3e**).

**Figure 3:**
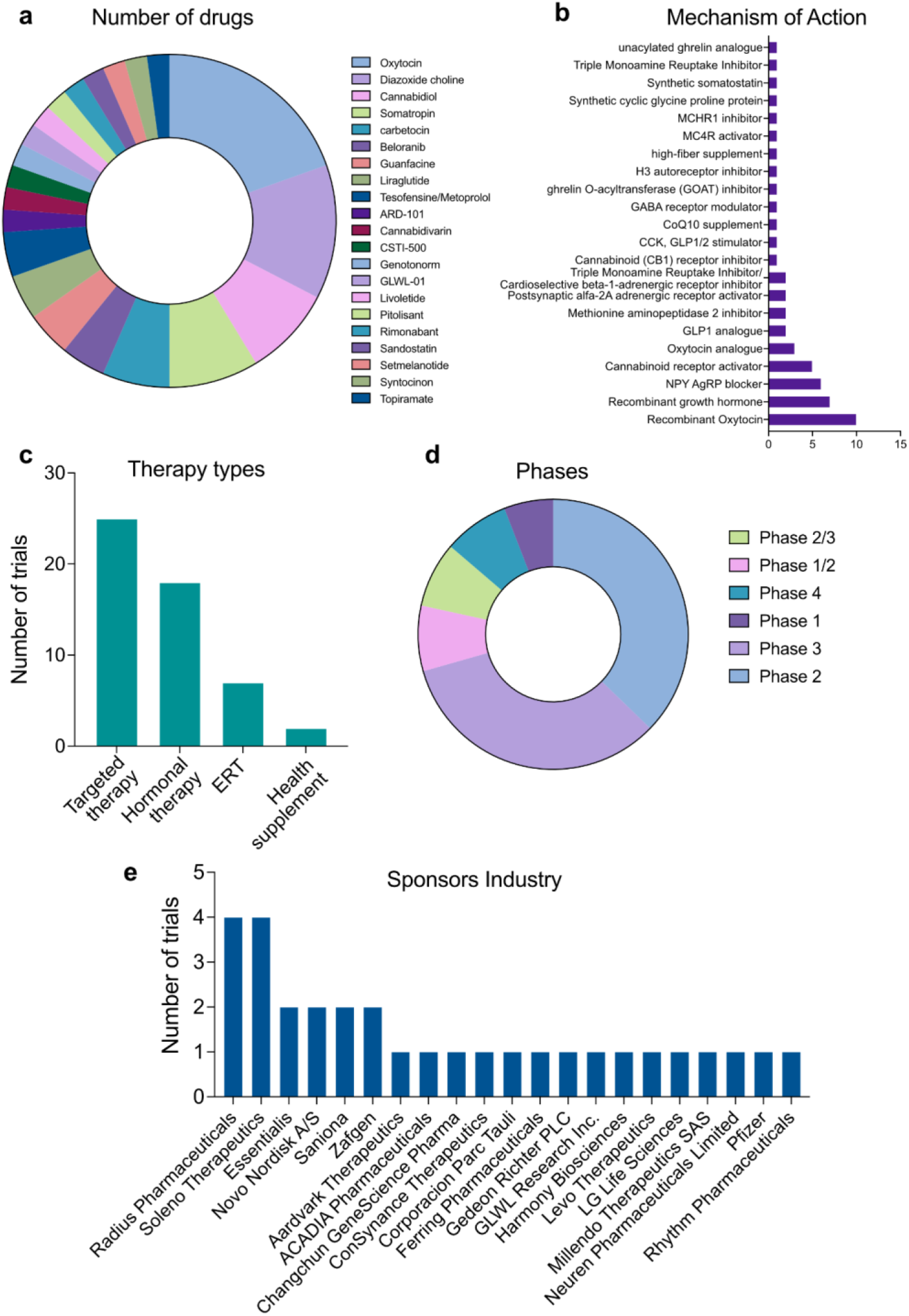
Clinical trial landscape in Prader Willi syndrome. (a) Pie chart showing the drugs in clinical trials against PWS. This representation covers “interventional” therapies. (b) Distribution of the key mechanisms of action (MoA) of the drugs against PWS. (c) Large sections of PWS therapies are targeted therapies. (d) Pie chart showing the phase distributions of the clinical trials against PWS. (e) A bar graph showing the major sponsors of the PWS trials.

### Efficacy landscaping: clinical endpoint analysis

PWS has a wide range of phenotypic manifestations among which hyperphagia and obesity is the most frequent and fatal feature having an early onset. There are a wide range of modalities used against PWS driven obesity and hyperphagia. There are instances of GLP1 usage against PWS patients, however there is no successful ongoing GLP1 mediated clinical trial. So we looked into the efficacy landscape of GLP1 agonists that has been reported to be used in PWS. This gives us a clear idea of the shortcomings, inefficacy results and adverse events landscape that served as the roadblocks for clinical studies. It also opens up opportunities for GLP1 agonist synergies with other molecules that can be tested as combinatorial therapy options against PWS driven obesity. Our second part of the analysis takes into clinical endpoint analysis and adverse events mapping of non GLP1 modalities covering the entire therapeutic landscape of obesity/hyperphagia targeted interventions.

### GLP1R agonist

Drug repositioning has emerged as an effective pipeline which can be used to twitch around on therapeutic targets common for different diseases. Another axis of repositioning pipeline is through analysis of correlated phenotypic features shared among several diseases. Prader Willi Syndrome manifests obesity and T2D as two of its prominent phenotypic features and thus shares similar molecular mechanisms. Lifestyle obesity and T2D have been treated with GLP1 agonists and have been established as the standard of care for lifestyle obesity. PWS manifests with obesity and T2D as two of its key features and GLP1 receptor agonists (GLP1 RAs) have been used for treatment in the form of case reports or clinical trials. The GLP1 RAs used in the studies against PWS are Dulaglutide (Trulicity) from Eli Lilly, Exenatide (Byetta, Bydureon) from Astrazeneca, Liraglutide (Saxenda and Victoza) from Novo Nordisk, Lixisenatide (Soliqua) from Sanofi and Semaglutide (Ozempic and Wegovy) from Novo Nordisk.

The following meta-analysis contains a detailed analysis of drug response from 33 PWS patients who were either adolescent or adults at the time of start of the GLP1 RA treatment regime. The drug efficacy landscaping of GLP1 RAs against PWS was performed using three primary clinical endpoints and one supplementary endpoint. The primary endpoints are (i) Percentage weight loss, (ii) BMI percentage decrease and post-treatment BMI and (iii) Glycated hemoglobin (HbA1c) decrease percentage in blood and post-treatment HbA1c concentration.

#### Percentage weight loss

Weight loss is a standard endpoint in clinical trials focused on reducing body weight. It typically denotes a decrease in body mass, quantified in kilograms or pounds, over a specified period. Accurate measurement is essential, often requiring standardized weighing conditions. While frequently the primary outcome in obesity trials, weight loss is primarily a surrogate endpoint for most health benefits. Consequently, additional endpoints are crucial to comprehensively assess an intervention’s clinical efficacy. Key factors in defining weight loss as an endpoint encompass measurement precision, timeframe, percentage change, and the minimum clinically important difference (MCID), representing the smallest weight reduction perceived as beneficial by patients. The weight loss percentage of all 33 patients were analyzed taking into account the change of weight between pre and post GLP1 RA treatment. The standard of care (SoC) for obesity, Wegovy, has a median weight loss percentage of 15% (**Figure 4a**). The responses to different GLP1 RAs can be segregated into three categories:

i. Category 1: Response comparable to SoC (**Figure 4a- i, ii**; black bars)
ii. Category 2: Response higher than SoC (**Figure 4a- i, ii**; red bars) and
iii. Category 3: Response lower than SoC (**Figure 4a- i, ii**; blue bars)

**Figure 4:**
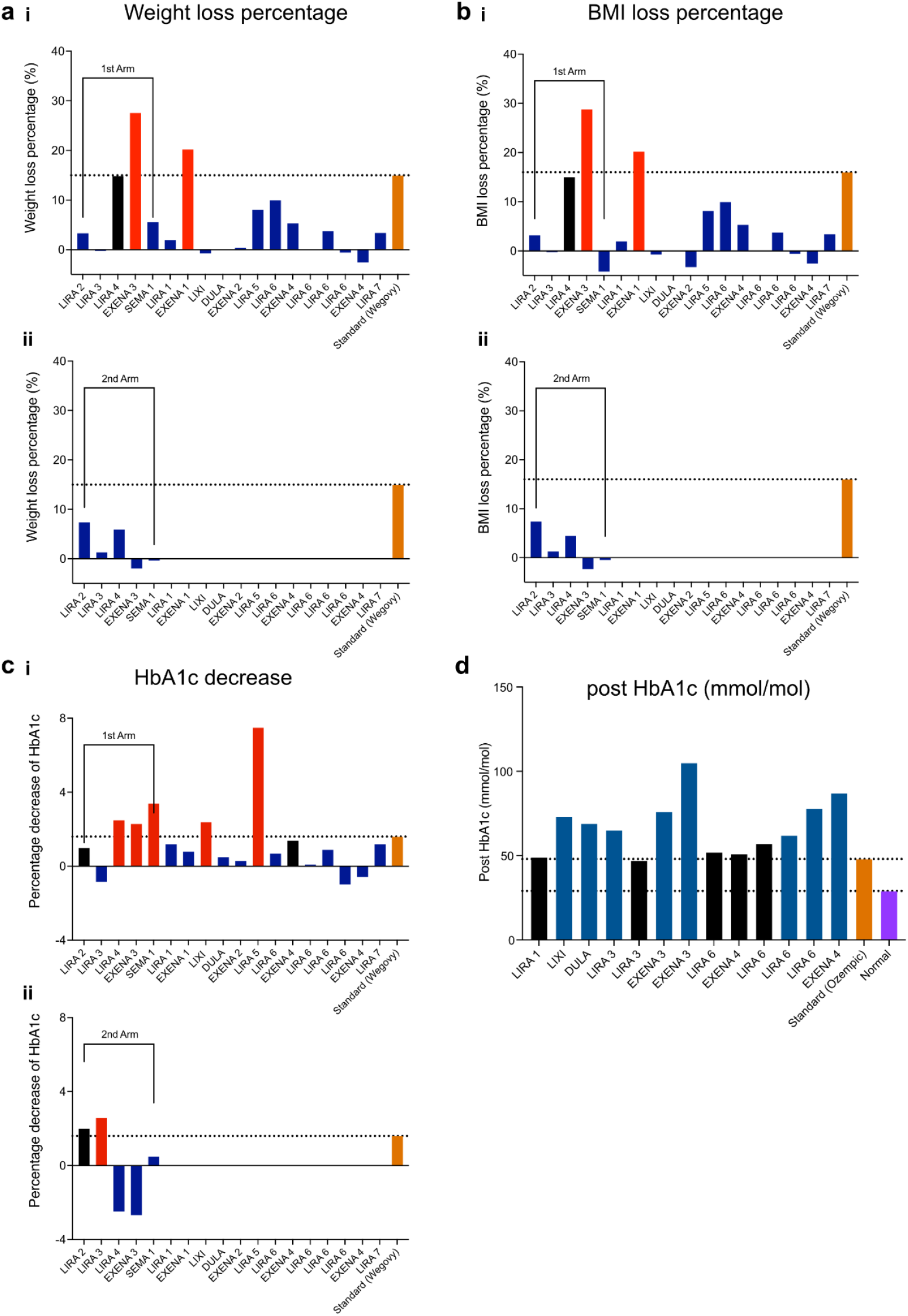
Clinical trial landscape in Prader Willi syndrome. (a) Meta analysis of weight loss percentage as a clinical endpoint for GLP1 RA in the (i) first arm and (ii) second arm of the treatments. Note that Wegovy (orange) is taken as standard of care at 15% weight loss. (b) Meta analysis of BMI loss percentage as a clinical endpoint for GLP1 RA in the (i) first arm and (ii) second arm of the treatments. Note that Wegovy (orange) is taken as standard of care at 15% BMI loss. (c) Meta analysis of HbA1c decrease as a clinical endpoint for GLP1 RA in the (i) first arm and (ii) second arm of the treatments. Note that Wegovy (orange) is taken as standard of care at 2% HbA1c decrease. (d) Post treatment HbA1c concentration across different GLP1 RA trials.

Category 1 comprises of liraglutide (Victoza) (**Figure 4a- i, ii**; LIRA_4), which showed comparable weight loss percentage to SoC in the first treatment arm of the study (1-6 months) (**Figure 4a-i**). However, it failed to show any clinical efficacy in the second treatment arm (6-12 months) (**Figure 4a -ii**). This study was from an extreme obese case where initial weight was 122 kg and height was 143 cm. Category 2 comprising of exenatide (Byetta/ Bydureon) showed increased weight loss percentage (5% and 12%) compared to SoC, in both the studies. In the first study (EXENA_1), it was found that out of the total weight loss, there was 9.2 kg of muscle loss and 6.6 kg of fat loss. Decrease in lean mass cannot be tolerated in any clinical studies and thus was discontinued (**Figure 4a-i**). In the second study (EXENA_3), there was an increase in weight loss percentage compared to SoC in the first treatment arm of the drug (1-6 months) (**Figure 4a-i**). However, it failed to show any clinical efficacy in the second arm of the treatment (6-12 months) (Figure 3b) for the patient who had extreme obesity conditions. Liraglutide (Saxenda), lixisenatide (Soliqua), dulaglutide (Trulicity), exenatide (Byetta/ Bydureon) and semaglutide (Ozempic) belongs to Category 3, which failed to show any significant improvement on weight loss percentage over treatment arms of 6, 12, 16, 24 months compared to the SoC (**Figure 4a-i**). All these individual case reports along with clinical studies suggest that the GLP1 receptor agonist is not completely effective in reaching the weight loss clinical endpoint for PWS driven obesity in comparison to lifestyle obesity SoC (**Figure 4a- i, ii**).

#### BMI percentage decrease and post-treatment BMI

BMI decrease and post-treatment BMI are key metrics in weight loss trials. BMI decrease quantifies the reduction in Body Mass Index from baseline to a specific follow-up point, serving as a primary or secondary endpoint. Post-treatment BMI represents the BMI measured after treatment completion, offering insights into the final weight status and potential weight regain. These parameters, while valuable, should be complemented by other assessments for a comprehensive evaluation of treatment efficacy.

The BMI percentage decrease of all 33 patients were analyzed taking into account the change of BMI between pre and post GLP1 RA treatment. The standard of care (SoC) for obesity (Wegovy) shows a median BMI percentage decrease of 15% (**Figure 4b- i, ii**; orange bar). The responses to different GLP1 RAs can also be differentiated into three categories

i. Category 1: Response comparable to SoC (**Figure 4b- i, ii**; black bars)
ii. Category 2: Response higher than SoC (**Figure 4b- i, ii**; red bars) and
iii. Category 3: Response lower than SoC (**Figure 4b- i, ii**; blue bars).

Liraglutide (Victoza; LIRA_4) in Category 1 showed a comparable BMI percentage decrease in comparison to SoC in the first treatment arm of the study (1-6 months) (**Figure 4b- i**). However, it failed to show any clinical efficacy in the second treatment arm (6-12 months) (**Figure 4b- ii**). This study was from an extreme obese case where initial weight was 122 kg and height was 143 cm (74). Exenatide (Byetta/ Bydureon) showed increased BMI decrease percentage (5% and 15%) compared to the SoC, in both the studies (**Figure 4b- i, ii**. In the first study (EXENA_1), it was found that despite efficient BMI decrease, there was 9.2 kg of muscle loss and 6.6 kg of fat loss upon weight segregation study. Decrease in lean mass cannot be tolerated in any clinical studies and thus was discontinued (75). In the second study (EXENA_3), there was an increase in BMI decrease percentage in the first treatment arm of the drug (1-6 months) (**Figure 4b- i**). However, it failed to show any clinical efficacy in the second arm of the treatment (6-12 months) (**Figure 4b- ii**) and was discontinued in the patient with extreme obesity conditions (76). liraglutide (Saxenda), lixisenatide (Soliqua), dulaglutide (Trulicity), exenatide (Byetta/ Bydureon) and semaglutide (Ozempic), belonging to the third category failed to show any significant improvement on BMI decrease percentage over treatment arms of 6, 12, 16, 24 months (77–83) in comparison with the SoC (**Figure 4b- i, ii**).

Post treatment BMI analysis reveals that more than 70% of the studies could not achieve the SoC level of post treatment BMI while some of them close to the SoC were mostly unchanged BMI from the pretreatment conditions. All these individual case reports along with clinical trial studies prove that the GLP1 receptor agonist is incapable of reaching the BMI decrease and target BMI clinical endpoint for PWS driven obesity in comparison to lifestyle obesity SoC (**Figure 4b**).

#### Glycated hemoglobin (HbA1c) decrease percentage in blood and post-treatment HbA1c concentration

HbA1c decrease in diabetes clinical trials refers to the reduction in glycated hemoglobin levels over a specified treatment period. HbA1c is a measure of average blood sugar levels over the past two to three months. Therefore, a decrease in HbA1c indicates improved blood sugar control in response to the intervention being studied. It is a critical endpoint in diabetes trials as it correlates strongly with the risk of diabetes-related complications.

The glycated hemoglobin (HbA1c) decrease percentage of 33 PWS patients were analyzed taking into account the change of HbA1c between pre and post GLP1 RA treatment. The standard of care (SoC) for T2D (Ozempic) (84) has a median HbA1c percentage decrease of around 2% (**Figure 4c-i, ii**; orange bar). The responses to different GLP1 RAs can be segregated into three categories:

i. Category 1: Response comparable to SoC (**Figure 4c- i, ii**; black bars)
ii. Category 2: Response higher than SoC (**Figure 4c- i, ii**; red bars) and
iii. Category 3: Response lower than SoC (**Figure 4c- i, ii**; blue bars).

Liraglutide (Victoza) and Exenatide (Byetta/ Bydureon) shows comparable Hb1Ac percentage decrease in PWS patients in comparison to the SoC against T2D (78) in both treatment arms (**Figure 4c- i, ii**). Lixisenatide (Soliqua), liraglutide (Victoza), exenatide (Byetta/ Bydureon) and semaglutide (Ozempic) belongs to the second category and shows higher HbA1c percentage decrease in PWS patients in comparison to the SoC in the first arm against T2D (74,75,79,80) in the first arm (0-6 months)( **Figure 4c- i**), but was inconsistent with the response in the second arm (6-12 months) (**Figure 4c- ii**). The third category comprising of liraglutide (Victoza), dulaglutide (Trulicity) and exenatide (Byetta/ Bydureon) shows lower HbA1c percentage decrease in PWS patients in comparison to the SoC against T2D in either of the clinical study arms (74,81,82) (**Figure 4c- i**).

Post treatment HbA1c concentration also reveals that almost 50% of the studies achieved the SoC level of post treatment (**Figure 4d**, black bars) while the rest of the GLP1 RAs showed higher HbA1c concentration compared to SoC (**Figure 4d**, orange bars).

The clinical endpoint analysis of HbA1c concentration in PWS patients reveals that GLP1 RA can have an impact on the T2D caused by PWS. The clinical efficacy of GLP1 RAs against PWS driven T2D, has a better physiological impact than that in PWS driven obesity (**Figure 4d**).

#### Liraglutide

Latest studies involving clinical trials on Liraglutide (Saxenda) from Novo Nordisk show that Saxenda fails to show any clinical efficacy against PWS driven obesity among 55 affected individuals. The clinical endpoints relating to BMI decrease showed no significant improvement upon treatment with Saxenda, a GLP1 RA. The change in mean BMI SDS (standard deviation score) was around −0.2/ −0.5 at week 16 and around -0.3/ -0.7 in 52 weeks in adolescents/ children respectively. The proportion of patients having more than 5% decrease in BMI was 23.2%- 34% in week 16 while it increased to only 30.7%- 33% in week 52 in adolescents/ children respectively. Thus it can be concluded that even if there was a profound difference in BMI change between the drug and placebo in the first treatment arm, the difference gradually narrowed down on the second treatment arm. In the second treatment arm, the proportion of patients having more than 5% BMI decrease did not increase significantly. Thus, it can be concluded that Liraglutide (Saxenda) is ineffective in PWS driven obesity management (73).

### Non-GLP 1 modalities

Therapeutic options targeting biomarkers other than GLP1 have been explored in the treatment of hyperphagia and obesity associated with PWS across various treatment regimens and clinical stages. A meta-analysis encompassing interventions from seven distinct therapeutic areas and one surgical procedure was conducted on a cohort of 771 patients. These interventions included: livoletide which is a UAG analog, oxytocin and its long-lasting stable variant carbetocin, diazoxide choline as a potassium channel inhibitor (AgRP-NPY blocker), setmelanotide as a MC4R agonist, beloranib as a methionine aminopeptidase 2 inhibitor, tesofensine as a triple monoamine reuptake inhibitor, and bariatric surgery (**Table 2**).

**Table 2:**
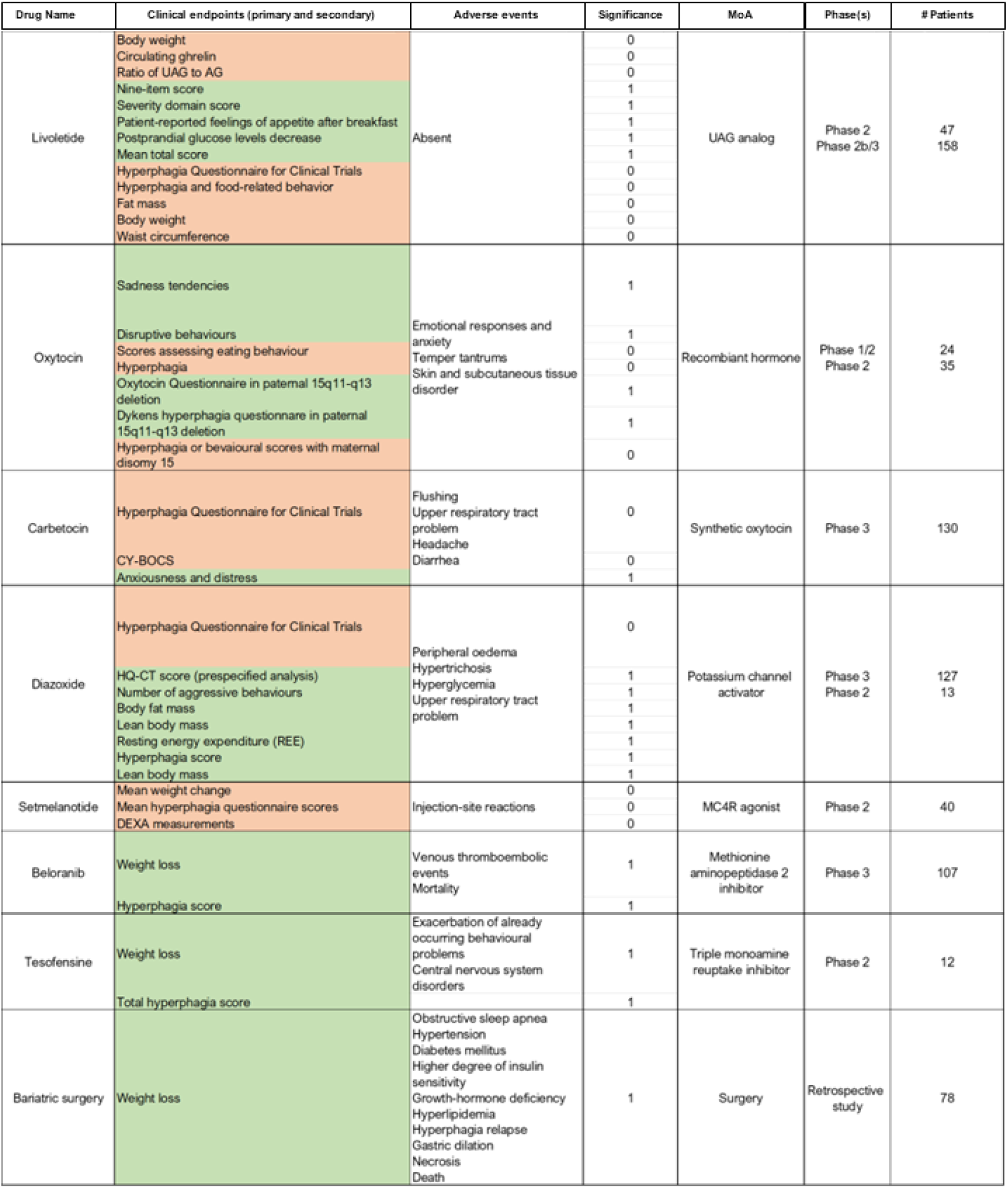
Clinical endpoints, adverse events and trial details for all non-GLP1 modalities explored in the treatment of hyperphagia and obesity associated with PWS.

Livoletide (n= 205; Phase 2, 2b/3) showed clinically significant changes in nine item score, severity domain score, patient reported feeling of appetite after breakfast, postprandial glucose level decrease and mean total score. However body weight, circulating ghrelin concentration, ratio of UAG to AG, hyperphagia questionnaire for clinical trials, hyperphagia and food related behavior, fat mass, body weight and waist circumference did not meet the statistical benchmark. No serious adverse events were recorded (85,86). Oxytocin (n=59; Phase 1, 2) led to significant improvements in sadness tendencies, disruptive behaviors, oxytocin questionnaire and Dykens hyperphagia questionnaire in paternal 15q11-q13 deletion. On the other hand, scores assessing eating behavior, hyperphagic behavior, hyperphagia or behavioral scores with maternal disomy 15 failed to reach significance. Sexual dimorphism between paternal and maternal PWS genotypes were observed in oxytocin response towards hyperphagia. The adverse events registered in these studies are emotional response and anxiety, skin and subcutaneous tissue disorder and temper tantrums leading to aggressive behavior (87,88). Carbetocin (n=130, Phase 3) treatment led to improvements in symptoms of hyperphagia along with anxiety and distress going down. The overall global impressions of change were recorded to be better. However, the hyperphagia questionnaire for clinical trials did not show any significant difference. Children’s yale-brown obsessive compulsive scale (CY-BOCS) score did not reach the significance benchmark. Some of the adverse events that occurred due to carbetocin treatment were diarrhea, flushing, headache and upper respiratory problems (89). Diazoxide choline (n=140, Phase 3) led to decrease in body mat mass, increase in lean mass and improvement of resting energy expenditure. A prespecified HQ-CT score analysis for patients with severe hyperphagia showed significance, but the overall HQ-CT clinical questionnaire was found to be insignificant. A phase 2 study showed improvement in hyperphagia scores but with low number of patients. This points to the fact that the effect of diazoxide choline on hyperphagia needs to be elucidated from a more mechanistic point of view. Some of the adverse events recorded were hyperglycemia, hypertrichosis, peripheral oedema and upper respiratory tract problems (90,91). Setmelanotide (n=40, Phase 2) did not meet the requirements of any of the clinical trials. Mean weight change, mean hyperphagia questionnaire scores and DEXA measurements showing lean mass gain all showed insignificant changes. Not many serious events were registered other than injection site reactions (92). Beloranib (n=119, Phase 2, 3) led to significant improvements in weight loss and hyperphagia scores with meeting a large proportion of the clinical endpoints. However this treatment led to severe adverse events like venous thromboembolic events and mortality of two (93). Thus it was discontinued. Tesofensine (n=12, Phase 2) showed significant outcomes in total hyperphagia scores and weight loss measurements. Some of the adverse events which were noted were CNS disorders and exacerbation of already existing behavioral problems (NCT03149445). However a previous study from 2008 shows tesofensine failed to induce any significant weight loss or improvement in patient quality of life (QoL) in a lifestyle obesity clinical trial study (94). Bariatric surgery (Retrospective study) leads to significant weight loss. However it has a huge spectrum of adverse events which makes it difficult. It causes relapse of hyperphagia, diabetes, gastric dilation, growth hormone deficiency, high insulin sensitivity, hyperlipidemia, hypertension, necrosis, obstructive sleep apnea and death (95,96). This meta-analysis elucidates that the majority of the analyzed treatments showed beneficial outcomes across multiple axes but they do not qualify for having a positive impact on the patients clinically. Neurological, obesity and QoL endpoints were found to be significant in the meta-analysis. However, none of the clinical trials analyzed achieved significance in all the primary clinical end points. A critical pain point that these clinical trials suggest is that hyperphagia is the most frequent primary endpoint which was not met. This infers that hyperphagia being the most manifested phenotype in PWS, should be studied in detail from a molecular point of view. Hyperphagic scales and measurement methodologies might need optimization and innovative addendums for better results.

Apart from the above medications which were discussed in details, there are several other approaches which have been employed to tackle the obesity/hyperphagia in PWS. The interventions target some of the key pathophysiology related to PWS. Orlistat, a pancreatic lipase inhibitor has proven to be effective in reducing fat absorption thus increasing lean mass. But, it has limited use in PWS due to poor compliance, gastrointestinal side effects, and modest efficacy (45,97). Metformin which is the first line of treatment for Type 2 diabetes and often used for insulin resistance may reduce appetite in some PWS individuals, but gastrointestinal issues can limit its use (98). Serotonin agonists like fenfluramine have shown short term efficacy in PWS patients. Sibutramine had the potential for reducing food intake, but was withdrawn due to cardiovascular risks (99,100). Lorcaserin induces at least 5% weight loss in obese patients and improves cardiovascular and diabetes risk factors compared to older serotonin drugs but lacks PWS-specific studies (101–103). Rimonabant, an endocannabinoid CB1 receptor inhibitor, is effective for weight loss but withdrawn due to severe psychiatric side effects (104). Bupropion, a sympathomimetic drug and naltrexone, an opioid receptor inhibitor, activates secretion of α-MSH, thus decreasing appetite and increasing resting expenditure energy. Naltrexone and bupropion independently target the brain’s reward system and hypothalamus and in combination, they appear to enhance weight loss by reducing appetite more effectively (105). Bupropion and Naltrexone show potential for reducing food intake and weight in obese patients, but require further study in PWS populations as a combinatorial therapy (106). Previous studies show that naltrexone as a monotherapy option, demonstrated modest benefits in body mass control, skin pricking and behavioral problems (107,108). Growth Hormone replacement therapy improves body composition and may delay obesity onset, but does not directly address appetite or food-seeking behaviors (27,109–111). While some of the above drugs show promise in addressing specific symptoms of PWS, there is a lack of effective, long-term treatments for the core symptoms of hyperphagia and obesity.

### Market analysis of drugs against PWS

A combination therapy involving standard of care drugs against obesity and T2D, as one of the major components of the therapy would require us to conduct and market research and commercial landscaping of the GLP1 RA drug space. The following section consists of:

1. Total revenue landscape of the obesity/ hyperphagia targeted therapeutics against PWS.
2. Obesity and Diabetes total market size and growth
3. Market share and growth trend of anti-diabetic GLP1 RA used against PWS
4. Anti-diabetic GLP 1 activators product specific EBITDA margin trend over 6 years (2018- 2023) used against PWS
5. Market share and growth trend of anti-obesity GLP1 RA used against PWS and
6. Anti-obesity GLP 1 activators product specific EBITDA margin trend over 6 years used against PWS (2018-2023).

#### Revenue landscape of obesity/hyperphagia targeted therapeutics against PWS

The obesity/ hyperphagia targeted drugs that have been studied against PWS show a very polarized distribution of the market shares. Wegovy, Ozempic and Trulicity are the highest grossing drugs with respective revenues of 13.9, 7.1 and 4.5 billion USD in 2023. Victoza and Saxenda are the precursors of Ozempic and Wegovy from Novo Nordisk which have turnovers of 1.2 and 1.5 billion USD respectively. Carbetocin, Oxytocin and Setmelanotide are some of the non-GLP1 modalities having negligible market size compared to its GLP1 counterparts (**Figure 5a**).

**Figure 5:**
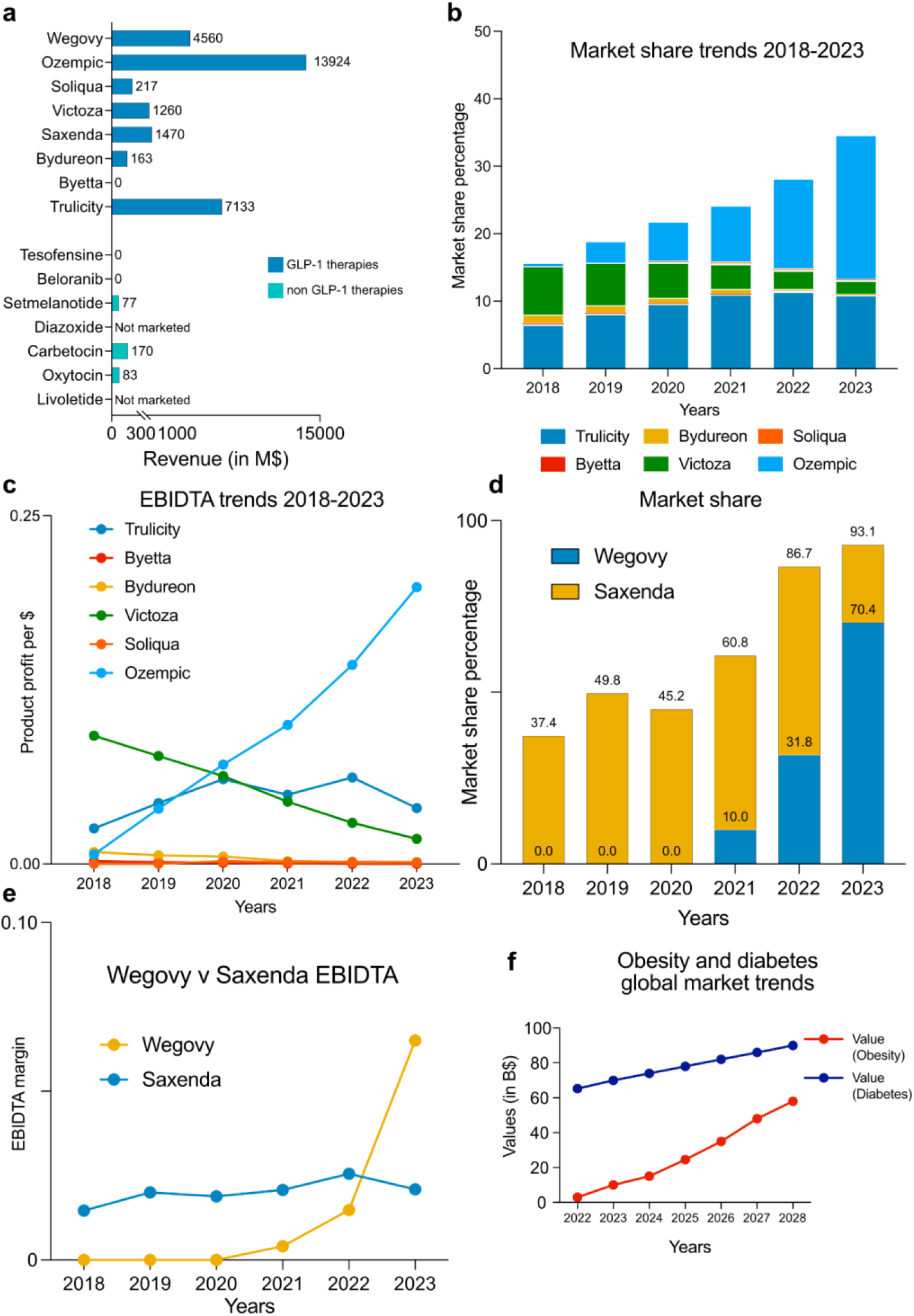
Market analysis of drugs against PWS. (a) Revenue in million dollars ($) of key drugs against PWS, dark blue indicate GLP1R agonists, while light blue indicates non GLP1 therapies. (b) Market share and growth trend of anti-diabetic GLP1 RA used against PWS (2018-2023). (c) EBIDTA trends of the key GLP1 RA described in panel b. (d) Market share and growth trend of anti-obesity GLP1 RA used against PWS (2018-2023). (e) EBIDTA trends of the key GLP1 RA described in panel d. (f) Diabetes and obesity total market size and growth.

#### Market share and growth trend of anti-diabetic GLP1 RA used against PWS (2018- 2023)

The anti-diabetic GLP1 RAs used for the PWS treatment were trulicity (dulaglutide) of Eli Lilly, byetta (exenatide)/ bydureon (exenatide) from Astrazeneca, victoza (liraglutide) from Novo Nordisk, soliqua (lixisenatide) from Sanofi and ozempic (semaglutide) from Novo Nordisk. The annual revenues of all the drugs were taken into account from 2018-2023 reported in the annual reports of Novo Nordisk, Eli Lilly, Astrazeneca and Sanofi. The individual revenue of each of the assets was expressed in ratio with the total market size of the diabetes market to calculate the market share of each of the drugs (**Figure 5b**). The analysis inferred the market share trend and the market growth of the anti-diabetic GLP1 RAs used against PWS. It was observed that in 2018, trulicity of Eli Lilly (**Figure 5b**; blue) and victoza (**Figure 5b**; green) of Novo Nordisk were the market leaders in terms of anti-diabetic GLP1 RAs drug space. Ozempic (**Figure 5b**; light blue) from Novo Nordisk had almost negligible market share as of 2018. In the course of 6 years (2018- 2022), Victoza lost its market shares drastically and Ozempic gained market shares from almost zero in 2018 to 21 % in 2023 clocking enormous positive growth. Trulicity maintained and experienced a decent increase in its total market shares and had a positive growth for 5 years (2018-2022). 2023 saw a marginal decrease in the total market shares of Trulicity. By the end of 2023 Ozempic is the market leader followed by Trulicity in the anti-diabetic GLP1 RA drug space. Byetta (Red) and Bydureon (**Figure 5b**; yellow) failed to achieve any market growth or gain of market shares in the drug space in 6 years (**Figure 5b**).

The product specific operational profitability (EBITDA margin) of each of the drugs was calculated taking into account their respective revenues, total revenue of the company and the company-wide EBITDA across the 6 years (2018-2023) (**Figure 5c**).

The EBITDA margin of each of the GLP1 RAs gives a picture of how much these drugs contributed to the total profitability of the company from 2018-2023. Victoza (**Figure 5c**; green) of Novo Nordisk had an EBITDA margin of almost 10% in 2018 which gradually decreased to around 1% in 2023. Trulicity (**Figure 5c**; blue) of Eli Lilly had an EBITDA margin close to 2% in 2018 which gradually increased and reached around 6% by 2022 followed by a drop to 4% in 2023. Ozempic (**Figure 5c**; cyan) of Novo Nordisk had a near zero EBITDA margin in 2018 and increased gradually to almost 20% by the end of 2023. However, Byetta (**Figure 5c**; red) and Bydureon (**Figure 5c**; orange) do not contribute significantly to the overall EBITDA margin of Astrazeneca and do not show any change or growth between 2018 and 2023.

#### Market share trend of anti-obesity GLP1 RAs used against PWS (2018-2023)

The potential anti-obesity GLP1 RAs used for the PWS treatment are saxenda (liraglutide) (**Figure 5d**; yellow) and wegovy (semaglutide) (**Figure 5d**; blue) from Novo Nordisk. The market share of these drugs were analyzed taking into account their respective revenue from 2018-2023, in ratio with the total market size of the obesity market. In 2018, Saxenda had almost 40% of the total obesity market shares which gradually increased to almost 60% in 2022. In 2023, the market share of Saxenda dropped to 23% due to the entry of Wegovy in the market. Wegovy was introduced in the market in June, 2021 and thus started to gain market shares from 2021 and had reached close to 30% overall market shares by 2022. In 2023, market shares of Wegovy catapulted to 70% making it the market leader in anti-obesity medication (**Figure 5d**). Saxenda of Novo Nordisk had an EBITDA margin of 1.5% in 2018 which increased to close to 3% in 2022 followed by a drop to 2% in 2023 (**Figure 5e**). Wegovy of Novo Nordisk had an EBITDA margin of zero in 2020 which gradually increased to around 1.5% in 2022 and shot up to 6% in 2023 making it one of the highest profitable assets (**Figure 5e**).

#### Diabetes and Obesity total market size and growth

Anti diabetes medication market reached a size of 65.3 billion USD in 2022 and is estimated to reach around 90 billion USD in 2028 with a CAGR of 5% (32) (**Figure 5f**). Anti-obesity medication market reached a size of around 3 billion USD in 2022 which is estimated to grow to 16 USD billion by 2024 and to around 58 billion USD by 2028 with a CAGR of 45% (33) (**Figure 5f**).

## Conclusion

Genetic obesity represents a complex and multifaceted health challenge. The condition’s strong genetic underpinnings significantly influence treatment outcomes, rendering traditional weight management strategies often insufficient. While lifestyle modifications remain essential, their impact is frequently limited due to the underlying genetic predisposition. Pharmacological interventions offer potential, but their long-term efficacy and safety require extensive investigation. Bariatric surgery, although a proven treatment for severe obesity, encounters challenges in the context of genetic obesity, as the condition’s heterogeneity complicates patient selection and surgical outcomes. Pharmacological agents have shown promise, yet their long-term safety and efficacy require further validation through extensive clinical trials. Bariatric surgery, although effective for morbid obesity, presents challenges in genetic obesity due to the variability in genetic backgrounds and associated comorbidities. Consequently, a personalized approach that considers an individual’s unique genetic profile is imperative. This necessitates a combination of strategies, including tailored dietary and exercise plans, potentially novel medications, and careful evaluation for surgical candidacy. Ultimately, achieving sustainable weight management in genetic obesity demands a comprehensive, interdisciplinary effort and ongoing research to unravel the complexities of this challenging condition.

The series of case studies, academic research and clinical trials involving GLP1 RAs against PWS driven obesity shows insignificant therapeutic efficacy. Thus, there still exists a high unmet need in the drug space against PWS where obesity and T2D are the major features that need to be targeted. However, the GLP1 RA is a standard of care against lifestyle driven obesity and Type 2 diabetes and thus can be combined with targeted therapies which are specific to PWS. Combination therapy emerges as a promising strategy, integrating GLP-1 RAs with PWS-specific treatments to optimize outcomes. Identifying appropriate combination partners and defining optimal dosing regimens are critical next steps. Moreover, a deeper understanding of the genetic and biological underpinnings of obesity is paramount for developing more targeted and effective therapies.

The obesity/ hyperphagia drug market exhibits substantial market share inequality. GLP-1 agonists, specifically Wegovy, Ozempic, and Trulicity, dominate the market, generating combined revenues of $25.5 billion in 2023. These drugs have eclipsed their predecessors, Victoza and Saxenda, also produced by Novo Nordisk, which generated $2.7 billion combined. In contrast, non-GLP-1 modalities like Carbetocin, Oxytocin, and Setmelanotide have minimal market presence. This stark disparity underscores the substantial clinical efficacy and market acceptance of GLP-1 agonists in addressing obesity and hyperphagia, particularly in comparison to other therapeutic approaches. The profitability of GLP-1 agonists, particularly Wegovy, has been exceptional. This high profitability is driven by several factors. Firstly, the significant unmet medical need for effective obesity treatments has created a robust market demand. Secondly, the drugs’ efficacy in promoting weight loss and improving associated health conditions has led to strong patient adherence and prescription rates. Thirdly, the relatively high cost of these medications contributes to substantial revenue generation. While Saxenda also experienced profitability growth, Wegovy’s rapid market penetration and superior efficacy have positioned it as a particularly lucrative asset. The sustained demand for effective obesity treatments suggests that the profitability of this drug class is likely to remain robust in the coming years.

The anti-obesity medication market is experiencing explosive growth, far outpacing the more established anti-diabetes market. While diabetes remains a critical health issue, the rapid rise in obesity rates has created a massive and rapidly expanding market for treatments. This dramatic shift presents a lucrative opportunity for pharmaceutical companies, driving significant investment in research and development focused on obesity therapeutics. As the market continues to expand, competition is likely to intensify, leading to advancements in treatment options and potentially lower costs for patients.

Long-term studies are essential to assess the safety and efficacy of emerging treatments in diverse genetic obesity populations. By addressing these areas, we can significantly improve the management of genetic obesity and enhance the quality of life for affected individuals. Future studies should focus on expanding our understanding of the genetic and biological mechanisms underlying obesity, which could lead to the development of more targeted and effective treatments. Furthermore, long-term studies are needed to evaluate the safety and efficacy of emerging pharmacological agents in diverse populations with genetic obesity. By addressing these areas, we can enhance the clinical management of genetic obesity and improve the quality of life for affected individuals.

## Data Availability

All data are available in the main text or the supplementary materials.

## Declaration of interests

All other authors declare they have no competing interests.

### Acknowledgements

We thank Emmanuelle Cacoub for reading the manuscript and providing critical feedback.

## Author Contributions Conceptualization

SS, MS Methodology: MS, NBL, DM Visualization: SS, MS Supervision: SS, HVH Writing—original draft: SS, MS

Writing—review & editing: SS, MS, HVH

## Data and materials availability

All data are available in the main text or the supplementary materials.

